# A systematic review and meta-synthesis of factors that influence clinical decision making for organ support interventions within the critical care unit

**DOI:** 10.1101/2024.12.07.24318649

**Authors:** Kenki Matsumoto, Brigitta Fazzini, Hannah Malcolm, Jack Eldridge, Zudin Puthucheary, Magda Osman, Timothy J Stephens

**Author notes:** Corresponding author: Timothy J Stephens.

## Abstract

**Background:** The critical care unit is a dynamic environment that necessitates a high volume of daily clinical decisions regarding organ support. It is known that decision-making varies significantly between clinicians, even where internationally accepted treatment guidance exists and overall the processes and influences on clinical decision-making are poorly understood. Our aim was to summarise the evidence on the decision-making process and the factors that influence organ support decisions in the critical care setting and 2) conduct a meta-synthesis to generate a model of medical decision-making, illustrating how different factors interact and affect the process.

**Methods:** We conducted a systematic search on three databases (PubMed, Embase and CINAHL) to find relevant papers exploring factors that influenced organ support decisions made by critical care clinicians. A meta-synthesis was then completed on included papers. The data were collated into a common format and cross-compared. This enable the generation of distinct themes/subthemes that were synthesised to develop a higher order interpretation.

**Results:** 33 studies (from 8967 citations) met the inclusion criteria. 21 of these only included nurses, 7 only doctors and 5 were interprofessional. 11 factors that influenced a clinician’s decision-making were identified: experience; professional and personal risk; uncertainty; characteristics of individuals; senior support; team hierarchy; decision making by colleagues; protocols, guidelines and evidence; time and workload; hospital structure; and clinical condition. These were grouped into four themes: human, team, system and patient factors. From our interpretation of the data, we found decision-making is often linear and primarily dictated by disease factors (i.e. patient’s clinical parameters). However, the identified human, team and system factors can place strain on decision-makers and make clinical scenarios more complex. There is scope however to modify these to optimise critical care decision-making.

**Conclusion:** While decision-making surrounding organ support is complex and dynamic, we identified recurring themes that influenced these decisions across different professions and environments. Further studies should focus on understanding how different decision-making processes directly affect patients’ outcomes.

## Introduction

Medical decision-making is a complex cognitive process, complicated by varying degrees of situational and clinical uncertainty (1). Historically, there were multiple competing theories of medical decision-making processes such as hypothetical deductive, intuitive and multiple processing models (2). However, the current prevailing model is a dual thinking process, whereby clinicians (indeed all humans) have two systems of decision-making (2, 3). System 1 thinking is typically described as low-effort, rapid and based on past experiences or heuristics whereas system 2 is a slower, more cognitively taxing process based on deliberation and reasoning. Even with this framework, medical decision-making can be highly variable from clinician to clinician even in the high-stakes setting of critical care. Examples include variation in decision-making and delivery of international accepted ventilation settings (4, 5), both in normal times and during the COVID-19 pandemic, and variation in sepsis treatment decisions (6, 7).

Critical care practitioners make over 100 decisions every day and often these are under significant pressure and time constrained manner. Additionally, the multidisciplinary nature of critical care entails a wide range of decision-makers differing in seniority and profession (8). This high degree of heterogeneity brings richness of opinion but may also negatively affect patient outcomes. Complex decisions around organ support treatments are varied and their outcomes can drastically differ (8). From a recent narrative review on decision-making around invasive mechanical ventilation and weaning emerged that nurse-led decisions are protocol driven compared to heterogeneous physician-led decisions which are guided by both subjective and objective information, and ‘gestalt’ (4). Another narrative review using decision-making theories to understand variation in critical care decision-making concluded that there was a need to increase the self-awareness of decision-making processes among critical care clinicians (2). This highlights the need to understand the decision-making process and the factors which affect clinicians at the individual and team level.

The aim of this systematic review and meta-synthesis is to address this knowledge gap by: 1) summarising the evidence on the decision-making process and modifiable factors around organ support decisions in the critical care setting and 2) meta-synthesis to create a higher order interpretation of the existing data, including generating the hypothesis for a potential new model of medical decision-making, illustrating how different factors interact and affect the process.

## Methodology

We reviewed literature from the period January 2002 to December 2021 in PubMed, Embase, CINAHL, following the Preferred Items for Systematic Reviews and Meta-Analyses (PRISMA) reporting guidelines and prospectively registered the review on PROSPERO (CRD42021283290).

### Search strategy and study selection

Search terms included “clinical decision making” OR “clinical decision making” OR “decision making” AND (“critical care” OR “Intensive care units OR “Critical care” OR ‘Intensive care units” OR “Intensive Care”, adapted for each specific database. A full list of search terms can be found in the Supplementary file. The search terms were made deliberately broad to capture all relevant studies. The initial search was followed by a forwards and backwards citation search. (9)

### Inclusion Criteria

All empirical studies on humans (of any age) that described or evaluated decision-making processes, and the factors that may influence these, on the commencement, titrating of and de-escalating of critical care interventions for organ support, including (but not limited to) ventilation, inotropes and renal replacement therapies. Qualitative, quantitative and mixed methods studies were all included.

### Exclusion criteria

- Not original research
- Not critical care specific
- Wrong outcome (not organ support decisions)
- Shared decision-making (i.e. making treatment decisions together with patients and their families)
- Admission or triage decisions
- End of life / withdrawal / palliation decision-making
- No access, not a full paper, not peer reviewed
- Not in English

### Combination of methodologies

An initial scope of the literature indicated that there was a mixture of methodologies in potential studies (quantitative, qualitative and mixed methods) and hence, a convergent integrated approach was planned (10). Extracted data from the quantitative studies and the quantitative component of the mixed method studies were transformed into qualitative data (“qualitising”) and subsequently a meta-synthesis conducted (11). This method was chosen as it allows the aggregation of existing data as well as the development of new concepts (12).

### Quality assessment

The Mixed Methods Appraisal Tool (MMAT) was used for quality assessment (13). This tool includes two screening questions followed by one of three distinct 5-point checklists for qualitative, quantitative and mixed methods studies. The quantitative and qualitative elements of mixed method studies were further assessed using the quantitative and qualitative checklists, respectively. As proposed, a cut-off score for quality was not set and the MMAT was used as a general guide of study quality. Two reviewers independently scored for inclusion/exclusion (KM / HM) with a third reviewer arbitrating in case of disagreements occurred (TS). Studies were excluded if they failed the first two screening questions of the MMAT and/or if there were sufficient concerns around the collection, synthesis or interpretation of the data. We acknowledge that variability in the quality of included studies may remain, despite removing lower quality papers which would not significantly contribute to the overall synthesis (14).

### Synthesis, qualitisation and data extraction

Established themes were compared using reciprocal translation and a “line of argument” synthesis was then conducted to determine how different factors interacted to influence a clinician’s overall decision-making (11). The Joanna Briggs Institute methodology was used to qualitise quantitative data (10). This was deemed preferable to quantifying qualitative data as it is less error-prone. An example of this is shown in Figure S1 in the supplementary file. Findings from quantitative papers and the quantitative elements from the mixed method studies were initially presented in its numerical format then transformed into textual descriptions. Extracted data was inputted into a common table. The distinction between first order constructs (participants’ descriptions often in the form of quotes) and second order constructs (original authors’ interpretations of the participants’ descriptions) often overlapped and therefore were synthesised together(15).

### Translation and translation synthesis

Quality variability has potential to affect theming, and the five higher quality papers based on the number of participants, methodology used and the MMAT score were themed first (13). The remaining 27 papers were subsequently themed, whilst observing for any emergent themes/subthemes. To examine how these different factors interact and develop a higher order interpretation (i.e. a “line of argument”), the translated themes and subthemes were continuously compared against each other alongside the original data. Emerging relationships between the themes/subthemes were merged to form a final line of argument synthesis (12, 16).

## Results of systematic review

8967 papers were identified via databases and citation searching. 141 studies progressed to full-text review, of which 33 were deemed eligible for inclusion (Figure 1). Of the 33 studies, 17 satisfied all points in the quality assessment, 13 had points where it was unclear whether there was sufficient information related to a criterion and three failed on one or two criteria in the 5-point checklist but it was felt inclusion was still justified.

**Figure 1:**
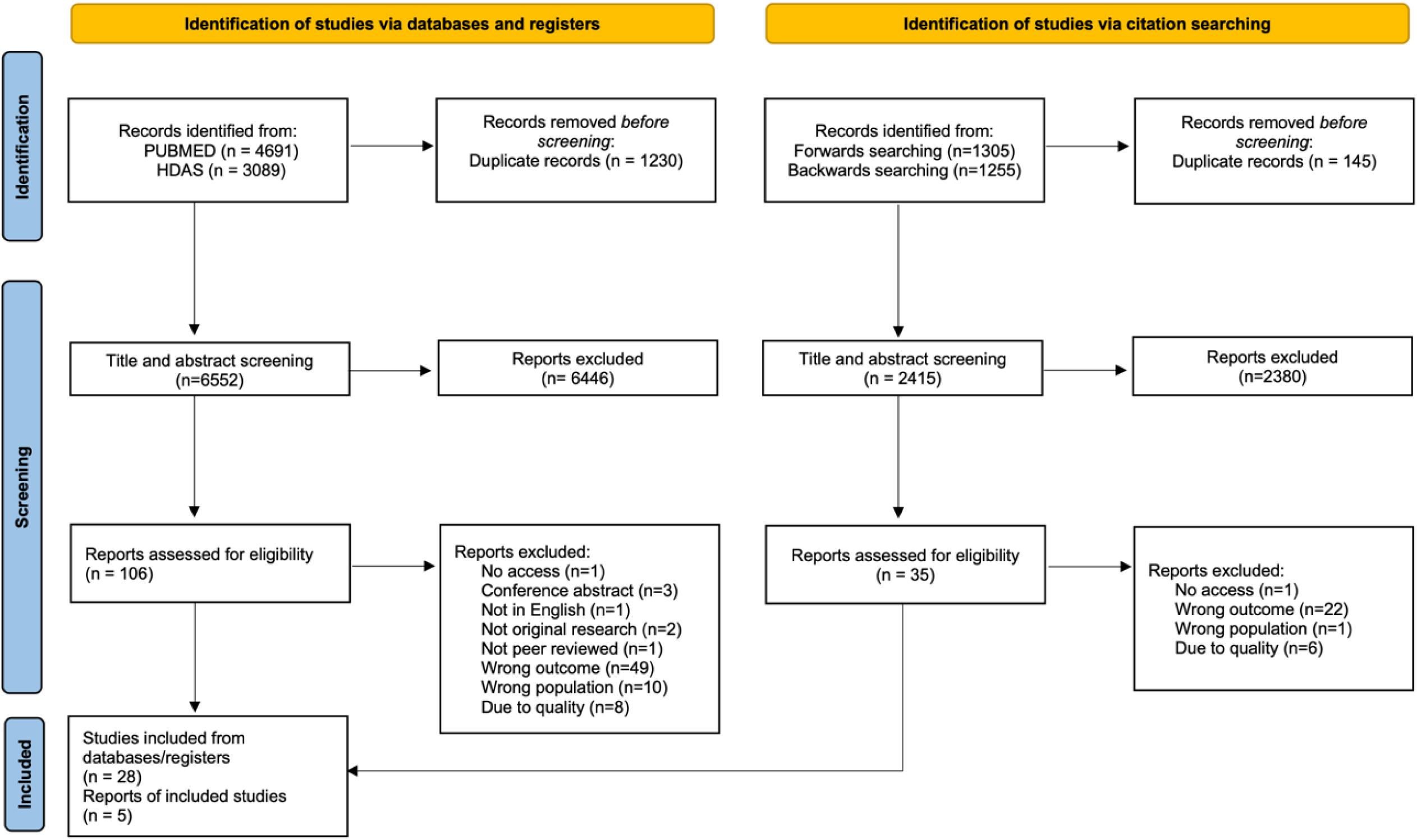
PRISMA flow diagram for the systematic review.

21 papers investigated decision-making in exclusively nurses, seven in exclusively doctors, five were interprofessional. 13 studies concerned decision making around ventilation and/or weaning, four on haemodynamic management, five on sedation, two on enteral feeding and nine around organ support generally. The majority of studies (22) were qualitative, four were mixed methods and seven were quantitative. The participant group seemed to affect the type of study – only five out of 12 studies in doctors used a qualitative methodology (four of these being interprofessional studies) compared to 21 out of 26 studies in nurses (see Table 1).

**Table 1.** Table highlighting the included studies in the systematic review, the professionals included in each study, the type of study and the decisions being assessed.

| **Title** | **Authors** | **Year** | **Professionals** | **Type of study** | **Decision** |
| --- | --- | --- | --- | --- | --- |
| Challenges and barriers to optimising sedation in intensive care: a qualitative study in eight Scottish intensive care units. | Kydonaki et al. | 2019 | Doctors, nurses and physios | Qualitative | Sedation |
| Clinical decision-making and mechanical ventilation in patients with respiratory failure due to an exacerbation of COPD. | Perrin et al | 2003 | Doctors | Quantitative | Ventilation and/or weaning |
| The clinical landscape of critical care: nurses' decision-making. | Bucknall | 2003 | Nurses | Qualitative | General organ support |
| An observational study of decision making by medical intensivists | McKenzie et al. | 2015 | Doctors | Quantitative | General organ support |
| Difficult to wean patients: cultural factors and their impact on weaning decision-making. | Kydonaki et al. | 2014 | Doctors and Nurses | Qualitative | Ventilation and/or weaning |
| Nurse decision making regarding the use of analgesics and sedatives in the pediatric cardiac ICU. | Staveski et al. | 2014 | Nurses | Qualitative | Sedation |
| Mechanical Ventilation, Weaning Practices, and Decision Making in European PICUs. | Tume et al. | 2017 | Doctors and Nurses | Quantitative | Ventilation and/or weaning |
| Factors influencing decision making in neonatology: inhaled nitric oxide in preterm infants. | Manja et al. | 2019 | Doctors | Mixed | Haemodynamic management |
| Dual agency in critical care nursing: Balancing responsibilities towards colleagues and patients. | Trapani et al. | 2016 | Nurses | Qualitative | General organ support |
| Expert clinical reasoning and pain assessment in mechanically ventilated patients: A descriptive study | Gerber et al. | 2015 | Nurses | Qualitative | Sedation |
| A study exploring factors which influence the decision to commence nurse-led weaning. | Gelsthorpe et al. | 2004 | Nurses | Qualitative | Ventilation and/or weaning |
| Protocolized weaning from mechanical ventilation: ICU physicians' views. | Blackwood et al. | 2004 | Doctors | Qualitative | Ventilation and/or weaning |
| Strategies and criteria for clinical decision making in critical care nurses: a qualitative study. | Ramezani-Badr et al. | 2009 | Nurses | Qualitative | General organ support |
| The factors which influence nurses when weaning patients from mechanical ventilation: findings from a qualitative study. | Lavelle et al. | 2011 | Nurses | Qualitative | Ventilation and/or weaning |
| Understanding nurses' decision-making when managing weaning from mechanical ventilation: a study of novice and experienced critical care nurses in Scotland and Greece. | Kydonaki et al. | 2016 | Nurses | Qualitative | Ventilation and/or weaning |
| The level of knowledge of respiratory physiology articulated by intensive care nurses to provide rationale for their clinical decision-making. | Pirret AM | 2007 | Nurses | Mixed | Ventilation and/or weaning |
| Critical care nurses' use of decision-making strategies. | Aitken LM | 2003 | Nurses | Qualitative | Haemodynamic management |
| Making decisions about medications in critically ill children: a survey of Canadian pediatric critical care clinicians. | Duffett et al. | 2015 | Doctors | Quantitative | General organ support |
| Ethical aspects of time in intensive care decision making. | Seidlein et al. | 2020 | Doctors and nurses | Qualitative | General organ support |
| Weaning from mechanical ventilation: factors that influence intensive care nurses' decision-making. | Tingsvik et al. | 2015 | Nurses | Qualitative | Ventilation and/or weaning |
| Nurses' near-decision-making process of postoperative patients' cardiosurgical weaning and extubation in an Italian environment. | Villa et al. | 2012 | Nurses | Qualitative | Ventilation and/or weaning |
| A comparison of novice and expert nurses' cue collection during clinical decision-making: verbal protocol analysis. | Hoffman et al. | 2009 | Nurses | Qualitative | Haemodynamic management |
| Preferred information sources for clinical decision making: critical care nurses' perceptions of information accessibility and usefulness. | Marshall et al. | 2011 | Nurses | Qualitative | Enteral feeding |
| A qualitative analysis of how advanced practice nurses use clinical decision support systems. | Weber S | 2007 | Nurses | Qualitative | General organ support |
| Why are physical restraints still in use? A qualitative descriptive study from Chinese critical care clinicians' perspectives | Cui N | 2021 | Doctors and nurses | Qualitative | Ventilation and/or weaning |
| Factors influencing nurse sedation practices with mechanically ventilated patients: a U.S. national survey | Guttormson | 2010 | Doctors | Quantitative | Sedation |
| Nurse staffing and unplanned extubation in the pediatric intensive care unit | Marcin | 2005 | Nurses | Quantitative | Ventilation and/or weaning |
| Experiences of intensive care nurses assessing sedation/agitation in critically ill patients | Weir | 2008 | Nurses | Qualitative | Sedation |
| The decision-making processes of nurses when extubating patients following cardiac surgery: an ethnographic study. | Hancock HC et al. | 2006 | Nurses | Qualitative | Ventilation and/or weaning |
| Clinical credibility and trustworthiness are key characteristics used to identify colleagues from whom to seek information | Marshall | 2013 | Nurses | Qualitative | Enteral feeding |
| The influence of patient complexity and nurses' experience on haemodynamic decision-making following cardiac surgery. | Currey et al. | 2006 | Nurses | Qualitative | Haemodynamic management |
| Intuition and the development of expertise in surgical ward and intensive care nurses. | King et al. | 2002 | Nurses | Qualitative | General organ support |
| ICU physician-based determinants of life-sustaining therapy during nights and weekends: French multicenter study from the Outcomerea Research Group. | Garrouste-Orgeas M et al. | 2014 | Doctors | Quantitative | General organ support |

### Description of themes

Theming of the five high-quality papers resulted in 12 distinct subthemes. Further thematic analyses (of the other included papers)(8, 17–43)) led to the generation of 20 subthemes that were subsequently revised and collapsed into 11 subthemes under four broader, high-level themes: human, team, system and disease factors.

#### Human factors

##### Clinician Experience

The dynamic environment in critical care units was reported as necessitating clinicians to absorb a high volume of moving data for clinical decision-making. There, however, are significant potential differences in both the assimilation and processing of this information across experience levels.

One such example is the use of cues. All clinicians use cues to aid decision-making ; however, experienced clinicians appear to identify cues that are higher in quantity and variety (44, 45). Less experienced clinicians report to encounter more cues before they have the confidence to enact their plans (28). This can result in experienced clinicians making proactive decisions compared to the reactive decisions made by less experienced clinicians (44). It was noted that clinicians often process information using heuristics or conscious pattern recognition (17, 20, 23, 24, 26, 34, 35, 38, 46). Experienced clinicians are reported to favour this type of information processing (20, 23, 24, 38, 46) while less experienced clinicians use this only in standardised tasks, such as nurse-led protocolised extubation (26). Heuristics and pattern recognition, however, introduce cognitive biases and partly explain why experience does not necessarily equate to better decision-making (17, 23). In fact, evidence suggests over-reliance on clinicians past experiences may lead to a lack of theoretical understanding to their own decision-making and/or inappropriately ignore existing evidence or guidelines.

##### Professional and Personal Risk

Critical care clinicians frequently make complex and high-stakes decisions that can have professional risk (47). Some clinicians seek to reduce the anxiety associated with this risk by altering their decision-making. One method reported in the literature is spreading the risk by discussing decisions with clinicians who are higher up the decision-making hierarchy, whether this is junior members of staff discussing with seniors (26, 48) or nurses and other allied health professionals discussing with doctors (23, 26, 48).

For example, a strategy described is reducing the number of decisions by refusing accountability for any decisions outside of the clinician’s perceived remit (35, 48). For example, decision-making around weaning is often driven by doctors, and there were reports of critical care nurses being reluctant to take accountability in weaning even when the decision making fell under their domain.

Clinicians may also choose to practice defensive medicine. Examples reported in the literature include clinicians ordering unnecessary investigations to anticipate issues before they arose (19) or try futile interventions in the hope of avoiding future litigation (46).

Another exacerbating factor may be specific team cultures, in particular a blaming culture (20, 39, 49) or one with a rigid hierarchy (39, 41). Studies also highlighted that level of risk that clinicians feel may be exacerbated by lack of support (26, 39, 48, 49). This could be from seniors (39, 49), other professions (e.g. nurses and doctors) (49) or the professional body (48). Conversely, a good leader can help alleviate this anxiety and optimise decision-making (26).

##### Uncertainty

Studies noted that uncertainty in decision-making may cause the clinician to slow down their thought process, but this can in turn may lead to delay in decision making and/or “freezing” (19, 36, 38, 46). Prognostication of critically ill patients, for example, can be especially challenging (36). Degrees of uncertainty mean that clinicians will often find it difficult to prognosticate the outcome of a patient and in turn whether to escalate or de-escalate treatment. Uncertainty may occur due to the complexity of the patient (19, 49) or the complexity of the decision (36), for example sedation management in patients with substance withdrawal syndromes or head injuries. It may also occur due to the clinician not previously facing a similar problem previously, especially if inexperienced (38). Due to this uncertainty, a clinician may delay making a decision until they gain more information (36).

##### Characteristics of the decision maker

There appeared to be wide variation in decision making amongst intensivists in the literature and part of this was due to underlying characteristics beyond solely their clinical experience (including age, gender, religious beliefs). The personality of the clinician, in particular their confidence, assertiveness and risk appetite, affected their willingness to make decisions (29, 38, 39). Clinicians who were younger and/or had religious beliefs were also found to be more likely to commence organ support (22) and female intensivists were found to make more decisions (8). Studies also outlined that a clinician could gain decision-making autonomy through formal education (28, 29). The role of the clinician appeared to be important but not their area of specialty (8, 33). Finally, clinicians interpret the same information differently and their view on a treatment can shape their decision-making around its utilisation.

#### System factors

##### Protocols and guidelines

Given the influences of uncertainty and perceived risk, one may expect guidelines and protocols to heavily influence decision-making. Instead, there was conflicting evidence on protocol and guideline utility regarding clinical decision-making. Interestingly, those studies advocating for protocol / guideline usage were from research contexts that did not have pre-existing local guidelines. These studies argued that implementing local guidelines would reduce variability (33, 38) and uncertainty in decision-making, and give a legal basis for decisions (48). Other studies that already had existing protocols argued that they were not helpful as they were often rigid, not customisable and not specific to the situation, especially when applying to a more complex case where the evidence is less clear (*e.g.* prolonged weaning) (21, 23, 28, 40, 41, 46, 48, 49). Instead, previous experiences (23, 29, 31, 41, 42, 45) or other clinicians’ experiences were preferred (31, 32). Some studies took a more nuanced view suggesting that protocols and guidelines are useful in certain situations. The weaning process, for example, could be protocolised as objective markers dominate in the decision-making process compared to the decision to wean, which is harder to quantify (23).

Protocols were also noted to be best used in conjunction with experience and therefore studies advised caution on inexperienced clinicians using these (18, 29, 42). On the one hand, it may help these clinicians work with confidence and within safe markers but on the other, it can result in decision-making without understanding the fundamentals leading to inflexibility in one’s decision making (18, 26, 29, 41). Ultimately, it was noted that implementation of protocols requires the support of the whole medical workforce, especially as clinicians prefer to obtain knowledge from colleagues over guidelines.

##### Time and workload

Due to the busy nature of critical care units, some decisions are made for pragmatic reasons rather than clinical ones. Time constraints and increased workload, for example, delays weaning or extubation decisions and extends sedation duration (26, 36, 38, 48). Staff levels have similar effects ; weaning was often only commenced when there was enough staff to complete observations and a lower nurse to patient ratio increases risk of unplanned extubation (25, 30). Conversely, higher nurse to patient ratios were associated with increased perceived autonomy and influence (40). The team composition was also of importance as a lack of experienced staff meant that these individuals spent more time supervising inexperienced staff members, thereby spending less time on their own decisions (19). Interestingly, having more patients appeared to increase the number of decisions made per patient (8).

##### Structure of hospital

The design and layout of the critical care may place a logistical strain on decision-making. It would be difficult, for example, to implement conservative measures for sleep promotion (*e.g.* through reducing noise and artificial lighting) if the critical care unit was too small and lacked daylight (20, 49). Bed shortages also has an anticipatory effect in decisions and may pressure early weaning for stable ventilated patients, although there is no evidence that it affects the provision of life-sustaining treatment (41). In units where clinicians were more fiscally aware, budgetary constraints and financial incentives also influenced decision-making (19, 36, 46). Ventilation, for example, was continued longer in units where this increased funding (36). Finally, corporate governance can have an impact on decision-making, with one study describing the unintended influence of increased physical restraint and sedation use in units where there was pressure to reduce unplanned extubation.

#### Team factors

##### Support

The level of support clinicians receives appeared to impact their decision-making, especially for those who are inexperienced. Studies found that without senior support, inexperienced clinicians find initiating management (e.g. reducing ventilator support) more difficult and make poorer decisions concerning complex patients (38, 45, 49). Conversely, senior support seemed to help clinicians detect issues earlier and respond more appropriately (45, 48). Formal education were also described to empower clinical decision-making (e.g. weaning) through providing not only the theoretical and practical skills (18, 48, 49) but also the credibility for the clinician to action their management plans (29).

##### Dependence on colleagues

Due to the collaborative nature of intensive care, decisions are highly interlinked and one decision can directly impact the next decision (19). If patients, for example, were not extubated in a timely manner, this could increase the risk of unplanned extubation and lead to use of physical restraints (20). Ambiguous communication was also described to impact decision-making. Studies reported that statements such as “wean off” or “wean as able” from more senior decision-makers can lead to different interpretations and more variable decision-making downstream (48, 49).

##### Team culture and hierarchy

Intra-professional and inter-professional hierarchy were also identified as important influences on decision-making. Those perceived to be at the bottom of the hierarchy generally had less autonomy to make decisions (26, 39, 41). This hierarchy was often determined by the clinician’s label (profession or grade) rather than the individual’s decision-making acumen.

Decision-making across the multidisciplinary team was more fluid in teams that harbour a collaborative atmosphere (23, 38, 48). Doctors, for example, may trust nurses to identify the patients who require weaning and instigate this. This arrangement, however, appeared dependent on the doctor’s trust on the nurse’s ability. Therefore, this delegation of decision-making authority was often reserved for experienced clinicians (18). Further, the ultimate decision-maker is still seen as those at the top of the hierarchy (18, 48). Senior nurses, for example, reported that they did not feel they could stop a weaning decision from a senior doctor (48). In other words, even in more collaborative teams, this implicit hierarchy on decision-making appeared to exist.

Beyond the multidisciplinary relationships, the unit-based practice and cultural norms also impact clinical decision-making (25, 26). Around 50% of nurses reported that other nurses’ knowledge and attitudes influenced their own sedation management (25). Further, clinicians (even experienced ones) adopt unit-based practice despite disagreeing with it.

#### Disease factors

The clinical condition of the patient and the associated disease factors were consistently perceived as the most important influence to decision-making (8, 20–24, 29, 30, 36–39, 49). The patient’s status alone could justify a clinician’s decision (18, 19, 35, 41). For example, a clinician may decide to work outside their perceived remit if there was clinical need (35). Experienced clinicians may also decide to take more control in complex weaning decisions (18, 41).

As well as the clinical scenario, the underlying patient co-morbidities heavily impact decision-making (23, 29, 49). Patients with mental health diagnoses are often kept sedated for longer (49) or patients with COPD are anticipated to need more complex weaning (29). Beyond the clinical characteristics of a patient, there are also explicit and implicit pressures from family (8, 36, 46). Some families, for example, may not respond well to mismatched expectations, such as the use of physical restraints to prevent unplanned extubation (20).

## Discussion

In this evidence synthesis we found that the patient’s clinical condition was the clearest theme (had the most codes contributing to it) across all papers. This resonates with a classic, linear process of clinical decision-making (figure 2a). There is a clinical scenario, dominated by the patients presenting condition, that the clinician interprets and based on this, clinicians decide upon their management plan. This commonly may involve heuristics, but also potentially more deliberate decision-making. If the decision was as simple as weaning ventilator settings according to the patient’s clinical parameters, the external influences on a clinical scenario may be limited. However, we also found key influences on the clinical scenario can place strain on the decision-making process. This may be because the content of the decision is more complex (e.g. initiating weaning from mechanical ventilation in a frail patient or difficult-to-sedate patients such as those with haemodynamic instability), multi-layered (e.g. requiring consideration of patient’s family or co-morbidity), the decision-maker is less experienced or there are other system influences (e.g. unit workload or hospital bed pressures) (Figure 2b). In these situations, there are external, influences that may affect the ‘linear’ decision-making process either as challenges or facilitators as outlined in Figure 2c. These are broadly split into human, system and team factors.

**Figure 2a.**
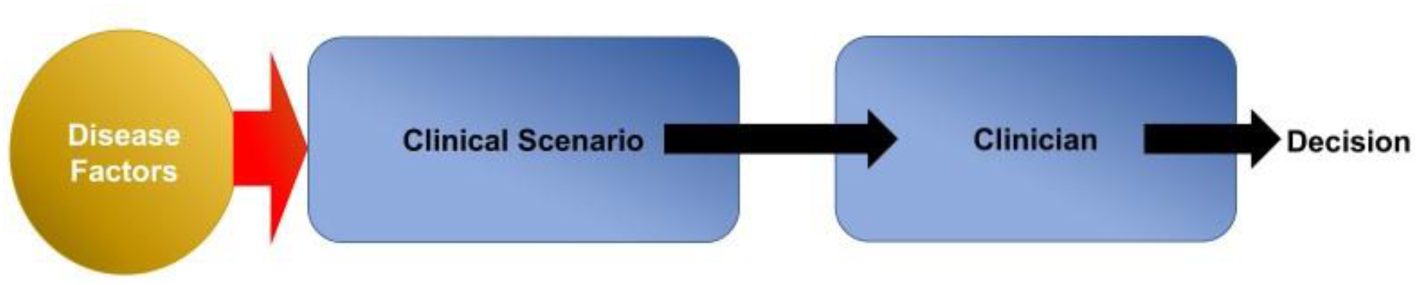
Linear decision-making, clinical scenario based on simple disease factors interpreted by the clinician leading to a decision.

**Figure 2b.**
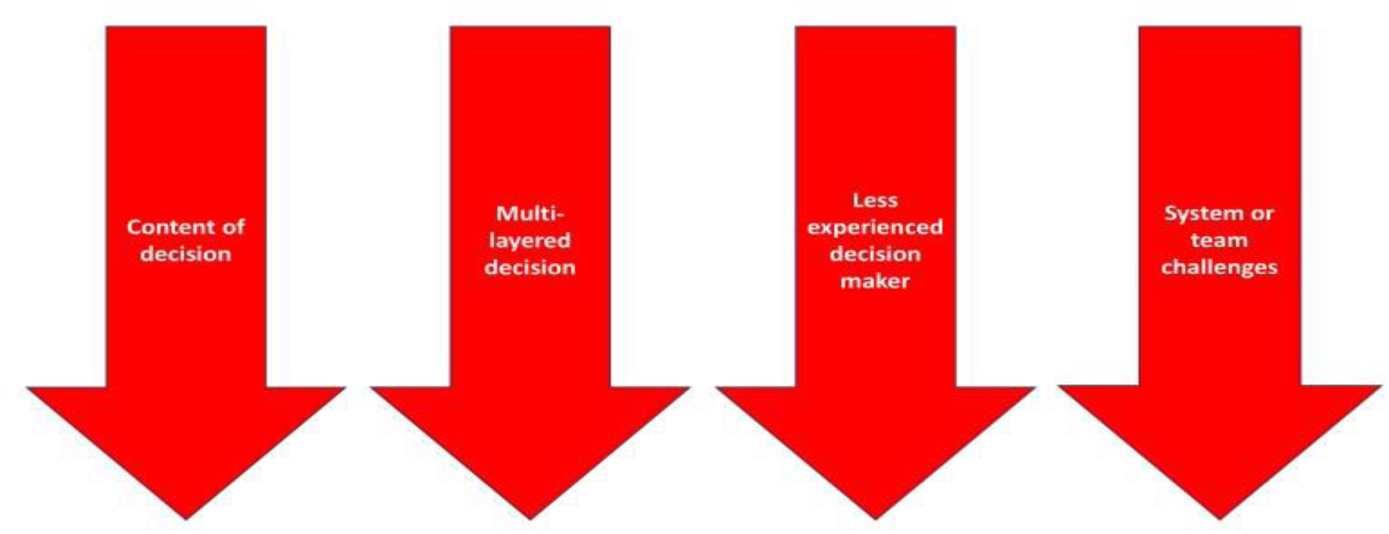
Examples of factors that switch the decision-making from a simple to a more complex model.

**Figure 3c.**
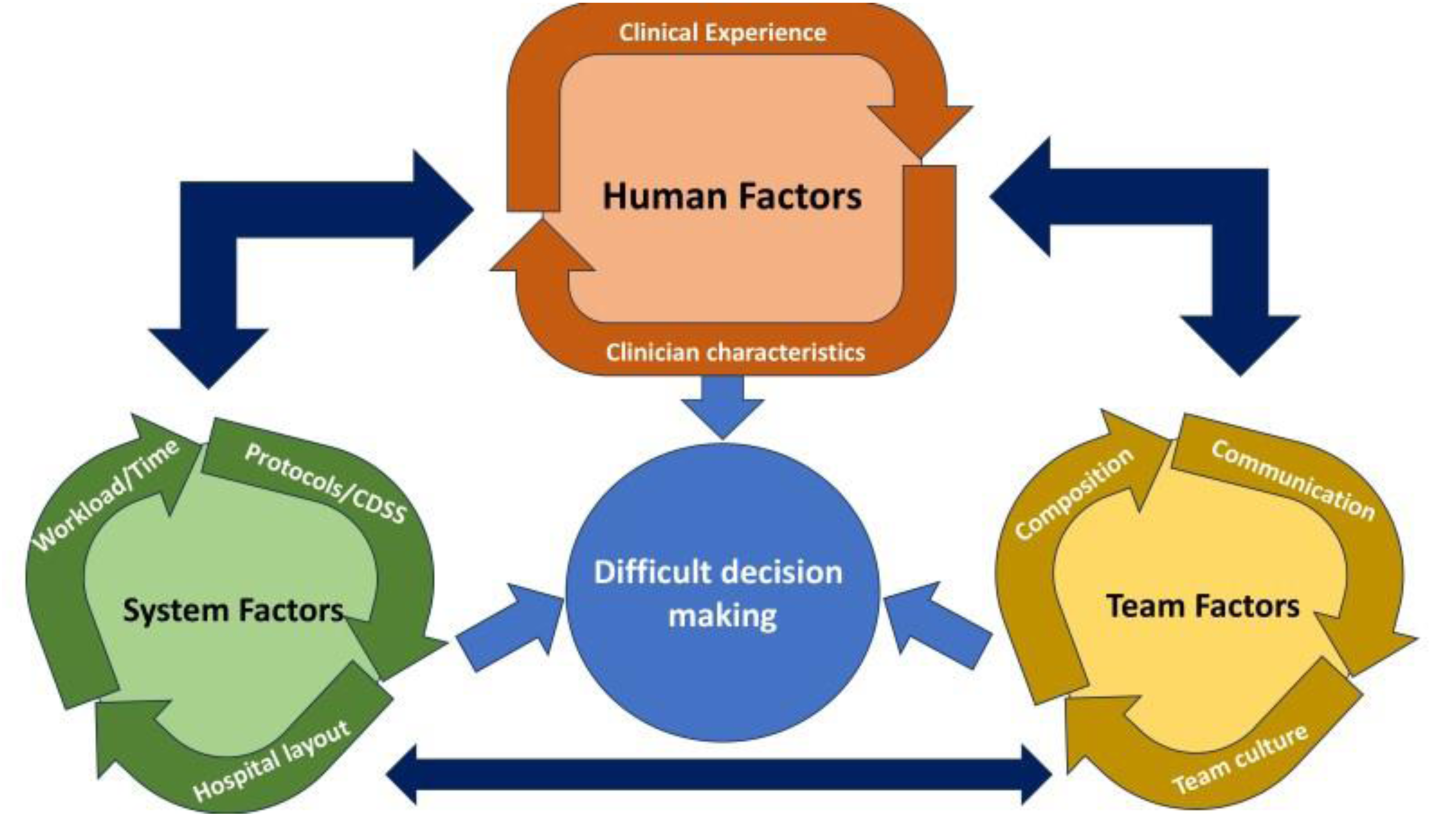
The different human, team and system factors that influence decision-making in difficult circumstances. Dark blue arrows highlight the interconnectivity between the different factors

### Human factor: the decision-making black box

In regard to human factors, our study has outlined the importance of experience in obtaining relevant, high-quality clinical parameters and in information processing. However, there was a lack of data around how experienced decision-makers navigated difficult decision-making. A recent study from our group investigating ventilatory decision-making in COVID-19 highlighted the importance clinicians attributed to anchoring past experience to their current problem (50). Those that believed COVID-19 presented similarly to previous cases of ARDS had higher confidence in their decision-making than those that did not. This suggests that intensivists are not comfortable in decision-making in uncertain conditions and the need to further study this. We however found in this review that there were fewer studies of decision-making in physicians (vs nurses) despite them often being the ultimate decision-makers. Further, the overwhelming majority of the existing literature in physicians was quantitative research rather than qualitative. This has meant that the decision-making process in intensive care physicians is still somewhat of a black box. Further studies utilising naturalistic observation are required to characterise the decision-making process and improve decision-making in difficult scenarios.

Our study also found that clinician’s characteristics has an influence on decision-making. However, most studies focussed on associating the characteristic with the decision made rather than the outcome of the decision (8). An experienced or confident decision maker can suffer from cognitive biases leading to poor decision making and so it is important for future studies to pair decisions with outcomes to understand which characteristics truly lead to improved decision-making.

### System factor: setting up the critical care unit for success

A recurring theme in this review was that the set-up of the hospital and critical care unit - in terms of its rota, workload and layout – can make easy decisions more difficult but conversely, difficult decisions more manageable (19, 26). This was no more evident than during the COVID pandemic where system fragilities were exposed. Decision-making was influenced by the lack resources, such as bed capacity, (50) and these system stresses directly impacted on clinician’s decision-making ability (50).

The literature remained equivocal in its opinion on protocol implementation in aiding decision-making. Real life data, on the other hand, highlights its utility. Data of ventilatory practices during COVID-19, for example, suggested that deviation from evidence-based ARDS management strategies led to worse outcomes (6). These contrary findings may be the result of the varying decisions made in the intensive care unit, some amenable to being protocolised and others less so. Decisions with objective parameters, such as the act of weaning, could be led by protocols but the decision to commence a wean may need a more complex clinical decision support system (CDSS) (4). The heterogeneity in the literature may also simply reflect the differing opinions of clinicians and a recent review highlighted the facilitators and barriers to the implementation of CDSS at the clinician, intervention and organisation level (4). Any future study looking to implement CDSS or protocols should not only consider the overall outcomes of its implementation but also survey clinician’s opinions prior to its implementations to determine whether there was a mismatch between clinician’s expectations to its outcomes.

### Team factor: creating a culture of support

Decisions within the intensive care unit relies on interprofessional teams with varying expertise (51). We found that collaboration and support in the unit increase each individual clinician’s autonomy in decision-making. This appears self-explanatory but this ethos is not implemented across all units. One barrier may be that delegation of decision-making requires a level of trust in an individual’s knowledge or skills with little or no shared history (51). This leads to a rigid hierarchy (both intraprofessional and interprofessional) and the sharing of decision-making powers only occur in those most experienced (18). In situ or multidisciplinary simulations may be a solution for understanding each member’s capabilities, as well as limitations and help improve team-based decision-making whilst protecting each clinician’s decision-making autonomy (52).

There is interconnectivity between human, system and team factors. Clinician experience affected its use of protocols/guidelines (human-system), the workload of an experienced clinician affected how well they could support a more junior team member (system-team) and a more supportive team allows clinicians to adjust the level of risk they are willing to take (team-human). In COVID-19, the most commonly co-expressed influences to decision-making were uncertain pathophysiology, limited resources and physical barriers, whilst strong teamwork was seen as a commonly identified mitigating factor (50). Further naturalistic observational studies are required to understand how these factors interact, especially in contexts where decision-making may be strained.

### Strengths and limitations

This study had several strengths. This systematic review identified nearly 9000 records and the inclusion and exclusion criteria were made *a priori* and were robust to answer our specific question on influences to decision-making surrounding organ support. Although the studies differed in quality, we used the MMAT to ensure that the synthesis was only on the highest quality papers. We also used an established method for aggregating the different methodologies used in the studies. Our findings, however, are limited to the available data and are prone to publication bias. For example, there were fewer studies, in particular qualitative ones, in physicians which may affect the generalisability of the study.

### Conclusion

Complex clinical decision-making is affected by four key factors, the human, the system, and the team as well as intrinsic disease factors. This study has generated the hypothesis of what the modifiable influences are, allowing us to understand how we can implement and leverage better decision-making. Further studies should focus on understanding how adjusting these modifiable influences to optimise decision-making process directly patients’ outcomes.

## Data Availability

All data produced in the present work are contained in the manuscript

