## Supplementary material for "A systematic review and meta-synthesis of factors that influence clinical decision making for organ support interventions within the critical care unit": Supplemenarty material

**Full search terms used**

**Search for PubMed:**

**Filters:** Papers published between 2001 - 2021, english language only

**Search terms:** ("clinical decision making"[MeSH Terms] OR “clinical decision making” OR “decision making”[Title/abstract]) AND ("critical care"[MeSH Terms] OR “Intensive care units[MeSH Terms] OR “Critical care”[Title/abstract] OR ‘Intensive care units”[Title/abstract] OR “Intensive Care”[Title/abstract])

**Search for EMBASE:**

**Filters:** Papers published between 2001 - 2021, english language only

**Search terms:** "((("critical care").ti,ab OR CRITICAL CARE/ OR INTENSIVE CARE UNITS/ OR ("intensive care units").ti,ab OR ("intensive care unit").ti,ab OR ("intensive care").ti,ab) AND (("clinical decision making").ti,ab OR CLINICAL DECISION MAKING/)) [DT 2001-2021]"

**Search for CINAHL:**

**Filters:** Papers published between 2001 - 2021, english language only

**Search terms:** "(("critical care").ti,ab OR CRITICAL CARE/ OR INTENSIVE CARE UNITS/ OR ("intensive care units").ti,ab OR ("intensive care unit").ti,ab OR ("intensive care").ti,ab) AND (("clinical decision making").ti,ab OR CLINICAL DECISION-MAKING/) [DT 2001-2021]"

“There are no significant group differences between respiratory and general physicians or intensivist in the decision to initiate ventilation. The variables studied were arranged in a similar order of important irrespective of specialty.”

The clinician’s area of specialty does not influence decision-making

Characteristics of the decision-maker

Quote from quantitative study

Transformation into a qualitative code (“Qualitising”)

The subtheme from this code

Figure S1: An example of “Qualitising” quantitative data
